# EVIDENCE FOR BIOLOGICAL AGE ACCELERATION AND TELOMERE SHORTENING IN COVID19 SURVIVORS

**DOI:** 10.1101/2021.04.23.21255973

**Authors:** Alessia Mongelli, Veronica Barbi, Michela Gottardi Zamperla, Sandra Atlante, Luana Forleo, Marialisa Nesta, Massimo Massetti, Alfredo Pontecorvi, Simona Nanni, Antonella Farsetti, Oronzo Catalano, Maurizio Bussotti, Laura Dalla Vecchia, Tiziana Bachetti, Fabio Martelli, Maria Teresa La Rovere, Carlo Gaetano

## Abstract

**Introduction & Background:** the SARS-CoV-2 infection determines the COVID19 syndrome characterized, in the worst cases, by severe respiratory distress, pulmonary and cardiac fibrosis, inflammatory cytokines release, and immunosuppression. This condition has led to the death of about 2.15% of the total infected world population so far. Among survivors, the presence of the so-called persistent post-COVID19 syndrome (PPCS) is a common finding. In patients who survived the SARS-CoV-2 infection, overt PPCS presents one or more symptoms such as fatigue, dyspnea, memory loss, sleep disorders, and difficulty concentrating. The pathophysiology of PPCS is currently poorly understood, and whether epigenetic mechanisms are involved in this process is unexplored.

**Methods & Results:** In this study, a cohort of 117 COVID19 survivors (post-COVID19) and 144 non-infected volunteers (COVID19-free) were analyzed using pyrosequencing of defined CpG islands previously identified as suitable for biological age determination. Besides, telomere length (TL) and ACE2 and DPP-4 receptor expression were determined. The results show a consistent biological age increase in the post-COVID19 population (58,44 ± 14,66 ChronoAge Vs. 67,18 ± 10,86 BioAge, P<0,0001), determining a DeltaAge acceleration of 10,45 ± 7,29 years (+5.25 years above range of normality) compared to 3,68 ± 8,17 years for the COVID19-free population (P<0,0001). A significant telomere shortening parallels this finding in the post-COVID19 cohort compared to COVID19-free subjects (post-COVID19 TL: 3,03 ± 2,39 Kb vs. COVID19-free: 10,67 ± 11,69 Kb; P<0,0001). Additionally, ACE2 expression was decreased in post-COVID19 patients compare to COVID19-free, while DPP-4 did not change.

**Conclusion:** In light of these observations, we hypothesize that some epigenetic alterations are associated with the post-COVID19 condition, particularly in the youngers (<60 years). Although the consequences of such modifications on the long-term clinical outcome remain unclear, this finding might help indicating a direction to investigate the pathophysiology at the onset of the persistent post-COVID19 syndrome.

## BACKGROUND

In the last trimester of 2019, a new severe acute respiratory syndrome (SARS) outbreak emerged due to a new Coronavirus (SARS-CoV-2) diffusion determining the COVID19 syndrome. At the time of this writing, the virus infected about 143 million worldwide, causing so far >3 million deaths (source: https://coronavirus.jhu.edu/map.html). More than 122 million survived the COVID19 syndrome (source: https://www.statista.com/statistics/1087466/covid19-cases-recoveries-deaths-worldwide/). Not surprisingly, nowadays, more and more frequently, survivors are coming to clinicians’ attention for the presence of post-COVID19 symptoms. Among others, the most commonly reported are fatigue, dyspnoea, memory loss, sleep disorders, and difficulty concentrating (1), which recently contributed to defining the so-called persistent post-COVID19 syndrome (PPCS) (1).

It has been observed that SARS-CoV-2 infected people who developed the adult respiratory distress syndrome (ARDS) also accumulated an excessive deposition of extracellular matrix in the alveolar compartment, causing pulmonary function’s worsening (2). Fibrosis accumulation has also been observed in the cardiovascular and nervous systems. The infection frequently caused increased circulating Troponin T and Brain Natriuretic Peptides, suggesting the presence of myocardium damage with possible activation of a remodeling process (3). Besides, a compromised heart function with reduced contractility has often been observed (4). Other effects were on the vascular system and included altering the coagulation cascade and clotting (5)(6). In this case, the fibrinogen pathway’s fast activation enhanced the risk of thrombotic events, leading to acute stroke and pulmonary embolism (7)(6). Interestingly, coagulation problems have been seen in post-COVID survivors and in PPCS patients, in which anticoagulants are routinely prescribed.

Similar to what was observed in 2003 SARS and the Middle East respiratory syndrome (MERS) epidemics, in COVID19 survivors, an increase of psychological disorders that include depression, posttraumatic stress, and anxiety has been observed (8). Similar disorders are often present in the general population exposed to stress determined by sociological factors often linked to economic damage or the sudden loss of some beloved ones (8). Remarkably, during the SARS or MERS epidemics, neuropsychiatric symptoms have been reported in hospital workers and caregivers (9). A similar situation has been reported in hospital workers assisting COVID19 patients (10).

Despite all the variety and significance of the symptoms reported by numerous COVID19 survivors with or without PPCS, valuable biomolecular markers to monitor this condition are still lacking.

Upon SARS-CoV-2 infection, the angiotensin-converting enzyme 2 (ACE2) expression level in the vascular system tends to decrease (11). This enzyme is involved in regulating the renine-angiotensin system (RAS); ACE2 contrasts the activity of the related angiotensin-converting enzyme (ACE) by converting the Angiotensin-II in Angiotensin (1-7) (12)(13). A low expression of ACE2 causes an accumulation of Angiotensin II, which may associate with exacerbating conditions leading to respiratory distress, hypertension, arrhythmia, cardiac hypertrophy, left ventricular function failure, atherosclerosis, and aortic aneurysms (13)(14). Moreover, ACE2 is negatively correlated with aging; it is relatively abundant in young and healthy people with significantly less risk of CVDs, while a lower quantity has been observed in the elderly (15).

The dipeptidyl-peptidase IV (DPP-4) is the receptor of MERS coronavirus (MERS-CoV) and has been reported in some cases to function as a co-receptor of SARS-CoV-2 (16). DPP4 expression increases on the surface of senescent cells (17), and its transmembrane form can cleave many molecules such as chemokines, neuropeptides, and incretin hormones. Because DPP-4 inactivates incretin hormones, high levels of DPP-4 have been observed in patients with type 2 diabetes mellitus and cardiovascular diseases. DPP-4 inhibitors have been used to treat T2DM, cardiac ischemia, and systolic dysfunction (18) (19). Some evidence suggests that DPP-4 inhibitors might reduce the coronavirus’s entrance into the airway, envisaging an additional therapeutic tool for COVID19 treatment (20). Whether the level of ACE2 and DPP4 in peripheral blood may represent valuable biomarkers to monitor recovery from COVID19 or the onset of PPCS is unclear.

In humans, telomere shortening is associated *in vivo* with the aging process, and, *in vitro*, it characterizes the cellular replicative senescence (21). Telomeres possess properties that make them suitable as biomarkers in several diseases or conditions, including cancer, CVDs, or aging (22) (23). The inverse correlation between telomere length (TL) and chronological age has been used for age prediction (24). Interestingly, among individuals infected by Sars-CoV-2, a reduced TL has been associated with the risk of developing more severe symptoms suggesting that TL at the moment of the infection might influence the clinical outcome (25). At present, little is known about the telomere dynamics during Sars-CoV-2 infection and in COVID19 survivors and whether this parameter might help predict the risk of developing PPCS.

In recent years, several studies aimed to identify biological or molecular markers of aging that correlate with chronological age and could be helpful to estimate the biological vs. chronological age (26). Some of these parameters have been defined based on modifications of the DNA methylome that correlate with the chronological age and might be used in age prediction models to define the biological age molecularly: the so-called DNAmAge (28). Many of these studies focalized on healthy or diseased individuals for forensic or public health problems (27) (28). Since the seminal Horvath’s work (29), several methods have been developed to estimate variation in methylation levels in selected DNA CpGs. These approaches apply to determining DNAmAge and have been used to emphasize the difference with chronological age: the so-called DeltaAge. Some methods are based on evaluating many CpGs, explored using a genome-wide array or next-generation sequencing technologies. However, others have been developed, taking into account a reduced number of CpGs analyzed by pyrosequencing (30)(31)(32). All systems are based on DNA methylation values obtained from whole blood samples. Because of their practicality, several pyrosequencing blood-based age-prediction models became recently popular, especially for forensic applications. These simplified methods have the additional advantage of being rapid and suitable to most laboratory settings without requiring bioinformatics (24)(33). A recent comparative study (34) analyzed the performance of some of these “reductionist” methods and found that the algorithm proposed by Bekaert B. et al., among others, worked well for biological age prediction in young and old subjects. This algorithm considers a prediction result correct for individuals aged 60 or higher when the predicted age matches the chronological age within a ±5.2 years range (24)(35). Considering that most post-COVID19 subjects fall within age 50 to 60 or higher, this method has been deemed suitable for the present study (24).

All the procedures to determine the epigenetic biological clock estimate DNAmAge and DeltaAge. A positive DeltaAge is considered an acceleration of the biological clock, while a negative DeltaAge indicates a bioage younger than the chronological one. In some studies, this parameter turned helpful to evaluate the risk of the onset of cardiovascular and neurodegenerative diseases, cancer, and the occurrence of death by all-cause (36).

In infectious diseases, the application of these methods is still limited. However, a DeltaAge acceleration has been observed in people infected by the Human Immunodeficiency Virus (HIV), the Cytomegalovirus, or bacteria such as the *Helicobacter pylori* (36). Besides, in post-mortem brain tissue, the DNAmAge of chronically HIV-positive individuals was higher than negative controls. Interestingly, a partial reversion of the accelerated DNAmAge has been recently observed following antiretroviral therapy (37)(38). Hence, HIV infection might change the epigenome landscape, potentially enhancing the risk of developing age-related diseases such as neurocognitive disorders (39). Similarly, in people infected by Cytomegalovirus, DNA methylation analyses performed in circulating leucocytes revealed an increased DeltaAge (40). The long-term consequences of these epigenomic alterations remain to be ascertained.

The present study investigates whether in COVID19 survivors there is a DNAmAge alteration and a DeltaAge acceleration, which, in association with other molecular parameters such as the telomere length and ACE2 expression in peripheral blood, might typify a set of biomarkers valuable in other and future studies exploring the risk of PPCS-associated pathophysiological manifestations.

## RESULTS

### Evaluation of DNAmAge and DeltaAge in COVID19 survivors

A cohort of 117 COVID19 survivors came to the attention of our physicians. Upon approval by the Ethical Committee and informed consent signing, peripheral blood was collected in EDTA vacutainers. A group of 144 age- and sex-matched COVID19-free volunteers with some risk factors partially overlapping with the post-COVID19 patients were recruited among the hospital workers and non-COVID19 patients (see Table I). Genomic DNA was extracted from the whole blood by a robotized station as described in methods. After bisulfite conversion and PCR amplification, pyrosequencing was performed. DNAmAge calculations were completed according to Bekaert et al. (24). Results indicate that the y-axis intercept differs significantly between the COVID19-free (Fig. 1A) and the post-COVID19 (Fig. 1B) population. The post-COVID19 group intercepted the y-axis at value 35.22, while the COVID19-free at 17.76. This difference determined an increment of DNAmAge of about 9 years in the post-COVID19 compared to the same group’s chronological age. No difference was appreciable in the controls (Table II). Accordingly, the vast majority (76.6%) of post-COVID19 had an average DeltaAge acceleration of 10.45 years (Figure 2, red dots). Considering that this method has a tolerance of about ±5.2 years (24)(35), the corrected average accelerated DeltaAge for this group was 5.25. On the contrary, all the COVID19-free volunteers had a DeltaAge of 3.68, falling well into the range of normality (24) (Figure 2, blue squares). The post-COVID19/COVID19-free DeltaAge ratio was 2.84 (Table II). Interestingly, the DeltaAge distribution within the two groups showed that the COVID19-free samples were evenly distributed between the normal (39,9%) and the accelerated range (48.9%) while the 12.8% had a decelerated biological clock. On the contrary, 76.6 % of post-COVID19 had an accelerated DeltaAge, with only the 23.4% falling in the normal or decelerated range (Figure 3A). Interestingly, while the COVID19-free DeltaAge was distributed evenly among the different age groups, the increase of DeltaAge in the post-COVID19 population was well represented among the younger people (age 56±12.8 years; Figure 3A, B). The older individuals, in both COVID19-free and COVID19-survivors groups, did not show signs of DeltaAge acceleration. This result indicates that the younger survivors might be more sensitive to the SARS-CoV-2-dependent remodeling of the epigenome landscape (Figure 3B).

**Table I:** Clinical information and risk factors in 117 COVID19 survivors and 144 COVID19-free volunteers.

| CLINICAL DATA | COVID19-free | Post-COVID19 |
| --- | --- | --- |
| Samples (n) | 144<br>(Male 66,0 %;<br>Female 34,0%) | 117<br>(Male 60,7%;<br>Female 39,3%) |
| BMI≥30 | 9,0% | 15,3% |
| Smokers | 37,5% | 16,9% |
| diabetics | 11,1% | 12,1% |
| Hypertension | 40,3% | 36,3% |
| Clinical history of CVDs | 33,3% | 27,4% |
| Antecedent lung involvement | 1,6% | 20,2% |
| COVID19-related complications |  |  |
| Pneumonia | / | 57,3% |
| Oxygen therapy | / | 52,4% |
| Artificial ventilation | / | 35,5% |
| Length of viral positivity (average) in weeks | / | 4,84 |

**Table II:**
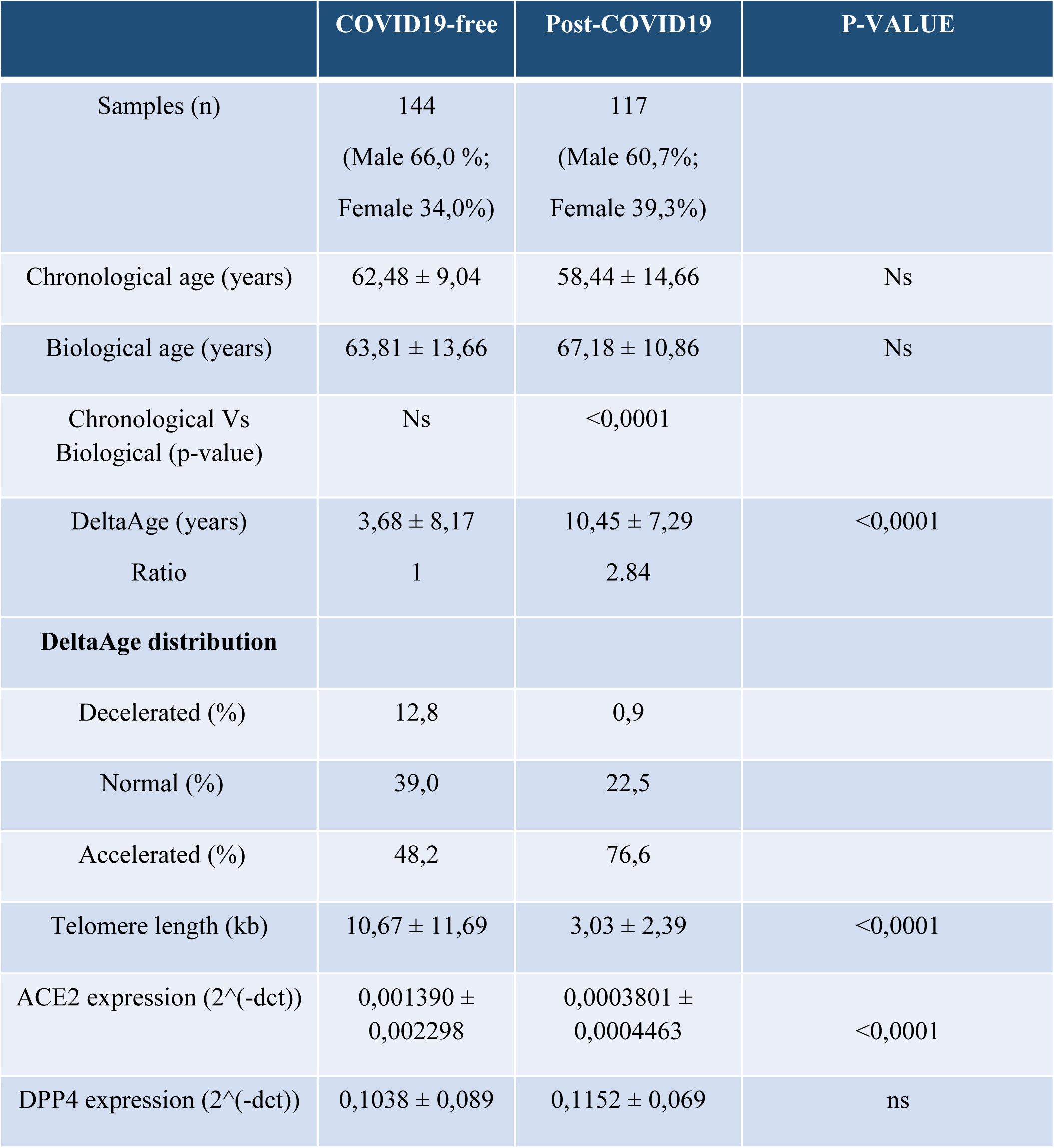
Summary

|  | COVID19-free | Post-COVID19 | P-VALUE |
| --- | --- | --- | --- |
| Samples (n) | 144<br>(Male 66,0 %;<br>Female 34,0%) | 117<br>(Male 60,7%;<br>Female 39,3%) |  |
| Chronological age (years) | 62,48 ± 9,04 | 58,44 ± 14,66 | Ns |
| Biological age (years) | 63,81 ± 13,66 | 67,18 ± 10,86 | Ns |
| Chronological Vs<br>Biological (p-value) | Ns | <0,0001 |  |
| DeltaAge (years) | 3,68 ± 8,17 | 10,45 ± 7,29 | <0,0001 |
| Ratio | 1 | 2.84 |  |
| **DeltaAge distribution** |  |  |  |
| Decelerated (%) | 12,8 | 0,9 |  |
| Normal (%) | 39,0 | 22,5 |  |
| Accelerated (%) | 48,2 | 76,6 |  |
| Telomere length (kb) | 10,67 ± 11,69 | 3,03 ± 2,39 | <0,0001 |
| ACE2 expression (2<sup>^</sup>(-dct)) | 0,001390 ±<br>0,002298 | 0,0003801 ±<br>0,0004463 | <0,0001 |
| DPP4 expression (2<sup>^</sup>(-dct)) | 0,1038 ± 0,089 | 0,1152 ± 0,069 | ns |

**Figure 1.**
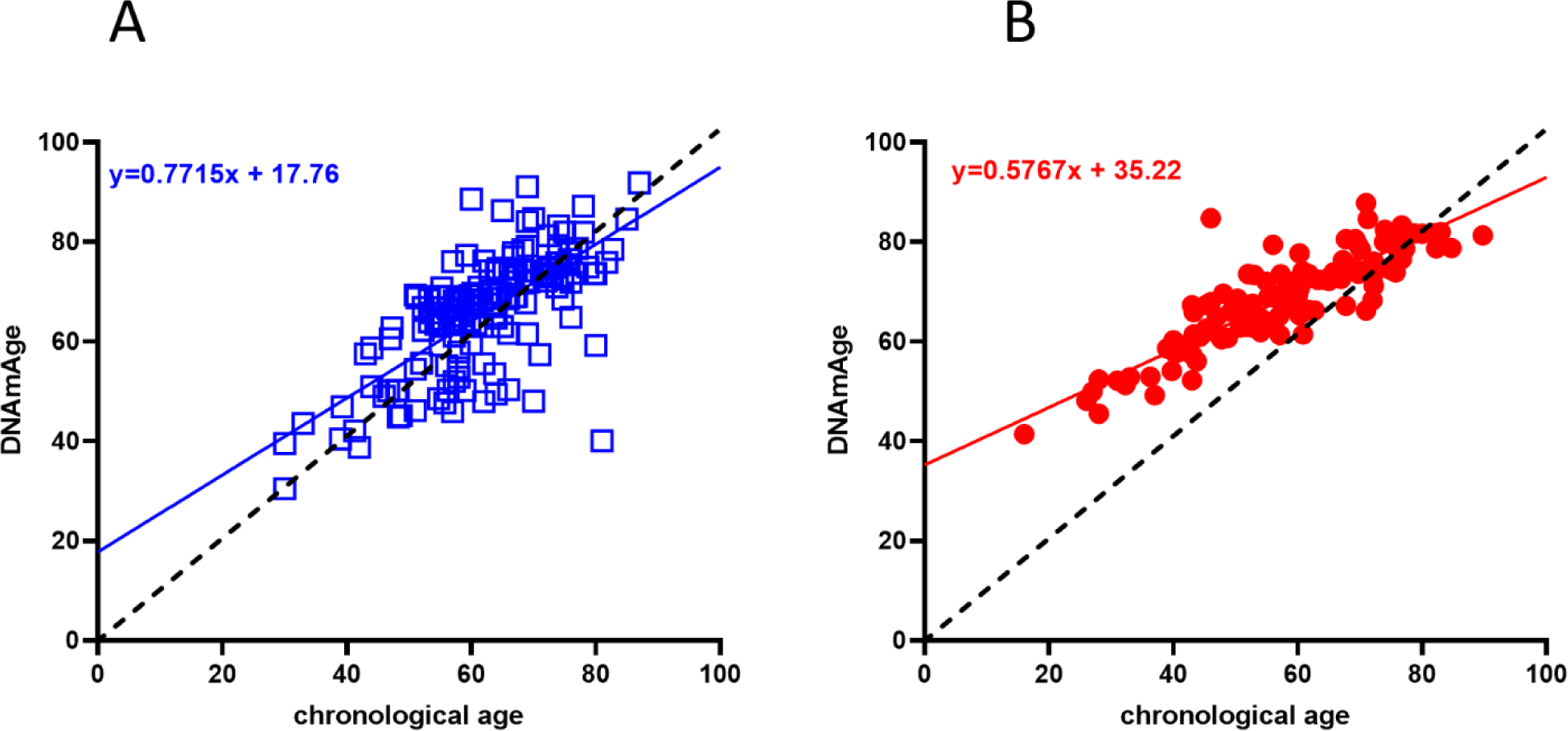
Biological age determination in COVID19-free (blue squares) vs. post-COVID19 (red dots). A) Linear regression of COVID19-free volunteers DNAmAge. B) Linear regression of DNAmAGE in the post-COVID19 subjects. In both graphs, the black dashed line is the bisector and represents the perfect correlation between chronological and biological age. The post-COVID19 group (right panel) showed a statistically significant DNAmAge acceleration.;P<0,0001 (two-sided T-test).

**Figure 2.**
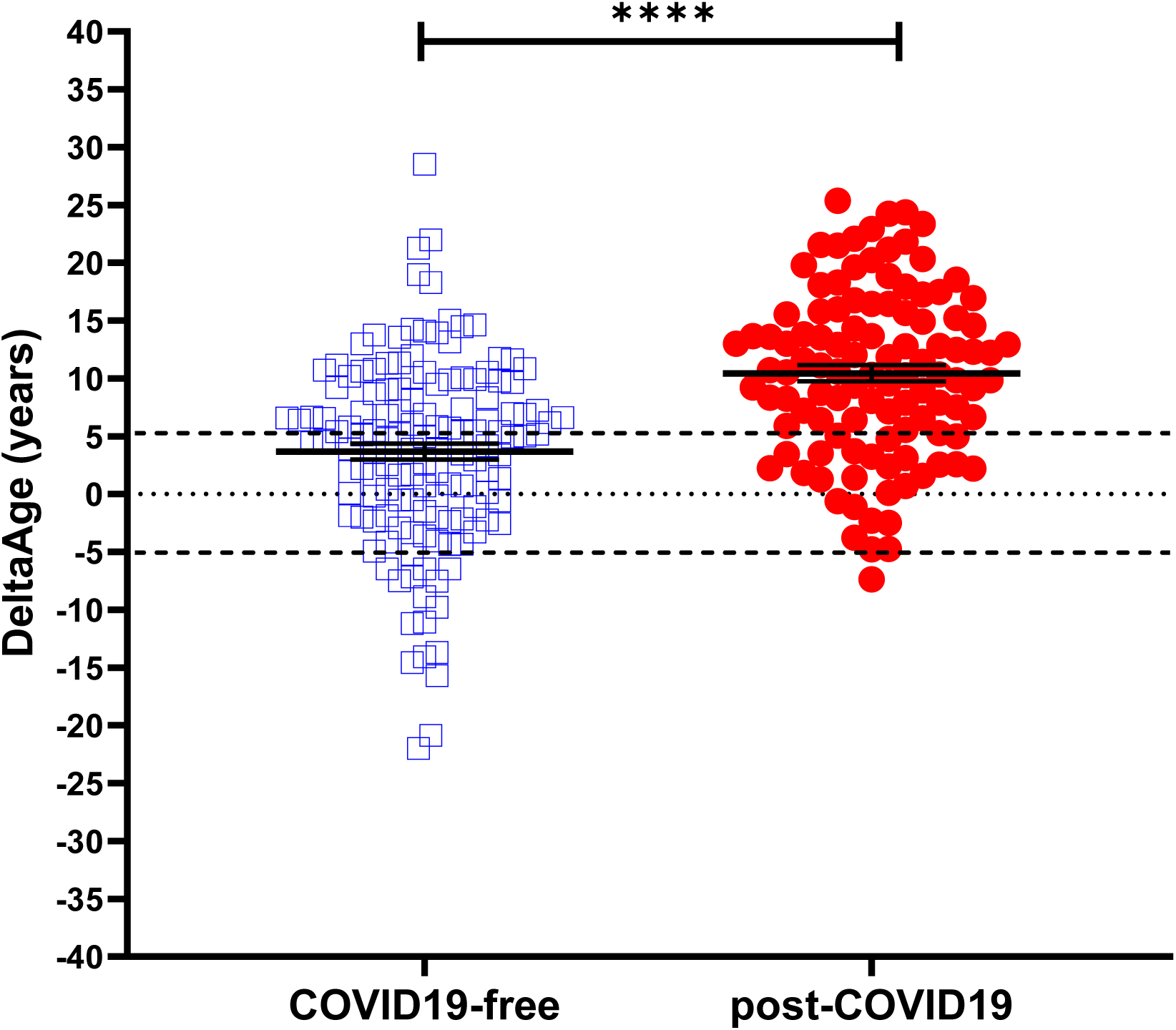
DeltaAge *distribution between COVID-free (left; blue squares) and post-COVID survivors (right; red dots). The black dashed lines indicate the ±*5 years *limit of the normal range according to the method. P*-value of <0,0001 (*Two sided T-test)*.

**Figure 3.**
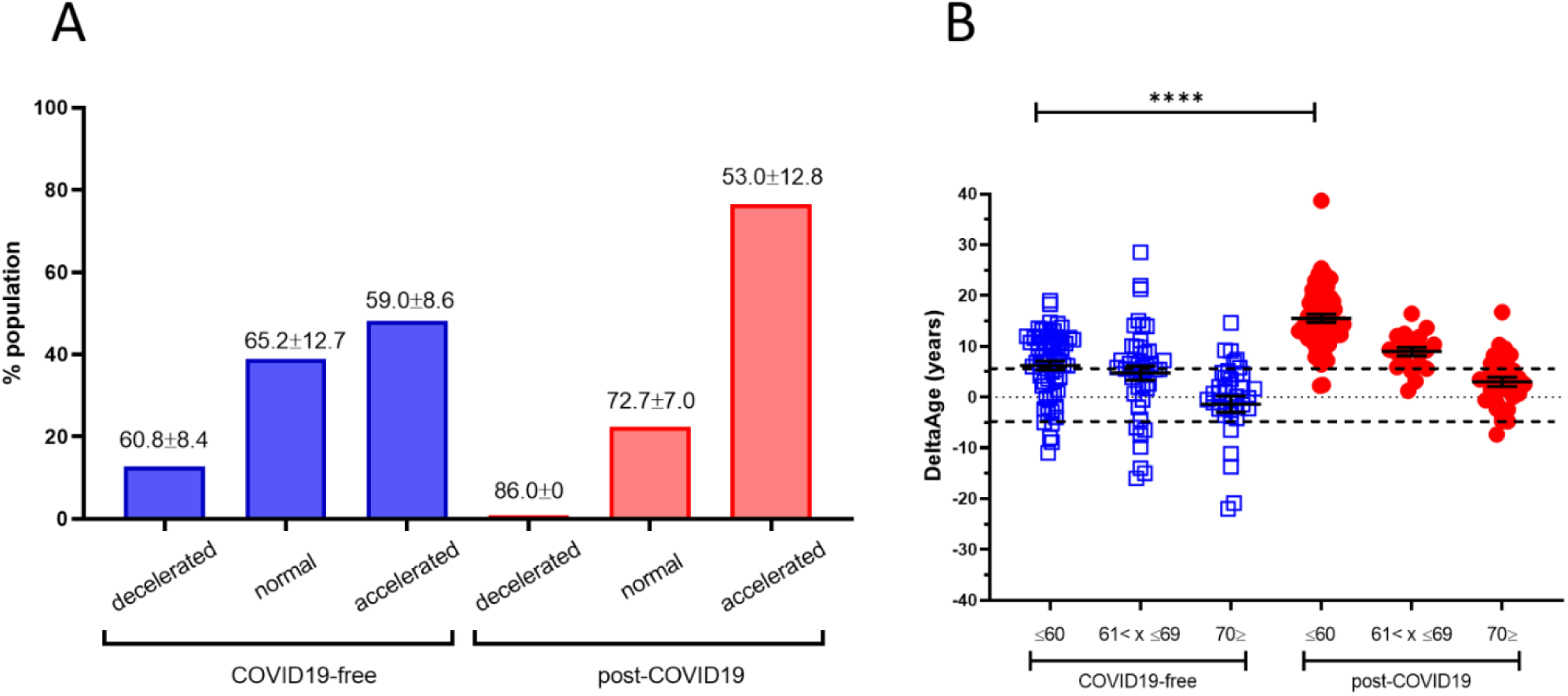
A) DeltaAge range of distribution within each age group. Specifically, in the COVID19-free cohort (blue bars), 12.8% of the participants were decelerated (mean:-8.7±5)*; 39.0% were falling within normality, while 48.2 % presented an accelerated DeltaAge (mean: +5.15± 4.34). Interestingly, only a negligible portion (0.9%) of post-COVID19 patients (red bars) are in the decelerated range. While 22.5 % were within the normal range, the vast majority (76.6 %) bore an accelerated bioclock (mean: +8.7 ±5.79). The average chronological age is reported above each bin. * DeltaAge mean values are considered after subtraction of the ±5.2 normality range distribution. B) The graph shows DeltaAge distribution according to the different chronological age groups. A significance of P<0.0001 between the younger COVID19-free group (<60) and the corresponding post-COVID19 patients is shown (Two-sided T-test)

### Telomere length quantification

TL shortening has been reported as a risk factor for developing more severe COVID19 syndrome (25). We investigated this parameter, which is also associated with the progression of the aging process (23). Comparing COVID19-free vs. post-COVID19 individuals revealed the presence of a significant chromosome ends shortness in the COVID19 survivors’ group (Figure 4A). Specifically, in the COVID19-free (Fig.4A; blue squares) volunteers, TL was 3.5 fold longer than in the post-COVID19 group (red dots). The correlation between DeltaAge distribution and TL indicates that post-COVID19 (Fig. 4C; red dots) compared with the COVID-free (Fig. 4B; blue squares) have shorter telomeres independently by an accelerated DeltaAge, suggesting that these two parameters might be regulated independently.

**Figure 4.**
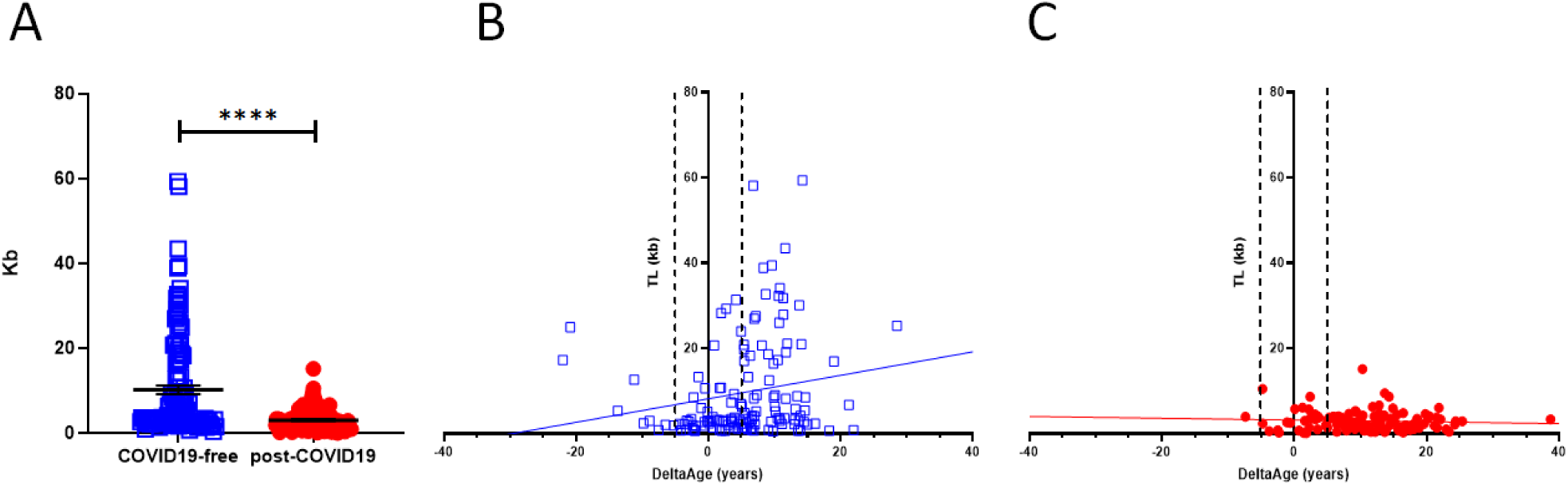
A) Telomere length analysis in COVID19 survivors (red dots) and COVID-free subjects. The graph shows that the COVID19-free group has longer chromosome ends compared to the post - COVID19. P-value <0,0001 (Two-sided T-test). B & C) Correlation between DeltaAge and TL in COVID19-free volunteers (B; blue squares) and post-COVID19 (C; red dots) patients.

### Peripheral blood expression of ACE2 and DPP-4

In a cell infected by SARS-Cov-2, the ACE2 expression decreases, but little is known about the intensity of this biomarker in post-COVID19 survivors. We evaluated the mRNA level of ACE2 (SARS-CoV and SARS-CoV-2 receptor) and DPP-4 (MERS-CoV receptor). The results are shown in Figures 5A and B. In the post-COVID19 population, at the time point in which the blood samples were taken, longer than four weeks from the end of the viral infection (see Table I), ACE2 expression was significantly reduced (Fig. 5A). The expression level of DPP4 was unchanged (Fig. 5B).

**Figure 5.**
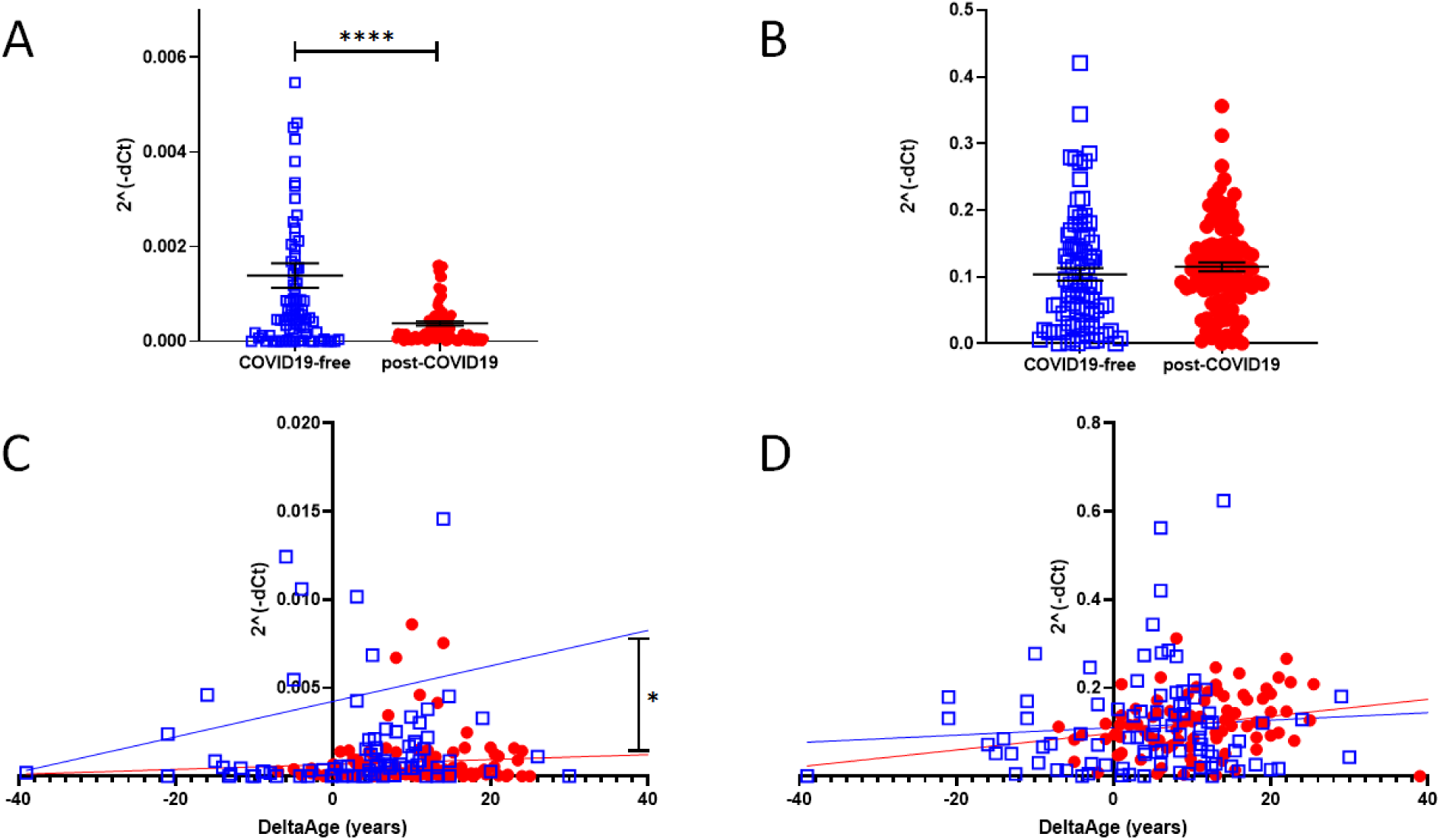
A) qPCR determination of ACE2 expression level in COVID19-free (blue squares) and post-COVID19 (Two-sided T-test: P<0.0001). B) mRNA level determination of DPP4 in COVID19-free vs. post-COVID19 showing no difference between the two groups. C) Correlation between DeltaAge and the relative expression levels of ACE2 mRNA in the peripheral blood of COVID19-free (blue squares) vs. post-COVID individuals (red dots). D) Correlation between DeltaAge and the relative expression levels of DPP4 mRNA in the peripheral blood of COVID19-free (blue squares) vs. post-COVID individuals (red dots).

The increased DeltaAge of the post-COVID19 group correlated well with the lowest ACE2 expression level (Fig. 5C; P<0.01). No differences were observed in the distribution of DPP4 by DeltaAge (Fig. 5D).

## Discussion

Nowadays, the global vaccination program against the SARS-CoV-2 is actively ongoing, and the incidence of COVID19 will soon decrease sensibly worldwide. Nevertheless, among the millions of COVID19 survivors, many will require long-term assistance due to increased post-COVID19 clinical sequelae defined as PPCS (1)(41). Despite the many manifestations associated with PPCS, there is a lack of molecular biomarkers potentially valuable monitoring PPCS onset and evolution. In this study, we took advantage of prior indication that biological age, defined as DNAmAge, could be altered in the presence of viral or bacterial infections (37)(40) (42) and from the fact that shorter telomeres have been associated with the risk of developing a worse COVID19 (25). In this light, we found that a consistently accelerated DeltaAge (+5.22 years above normal range) characterized the post-COVID19 population, and particularly those chronologically under 60 years (Figure 3A and B). This observation was parallelled by a significant telomere shortening (Figure 4). Although the two parameters seem independent (Fig. 4B and C), both alterations coexisted in the post-COVID population. The pathophysiology at the basis of these findings remains unclear; however, they may reflect a modified epigenetic environment, particularly evident among the younger COVID19 survivors (Fig.3). It is well known that the progression of aging is associated with critical metabolic changes. Some of these changes occur in the level of metabolites regulating the function of essential epigenetic enzymes, such as the decrease in NAD+ levels, the cofactor of Sirtuins (43), and the reduction in alpha-ketoglutaric acid (44), the cofactor for all dioxygenases, including those deputed to proteins and DNA demethylation (45). Although very speculative, it may be that older adults are relatively less sensitive to SARS-CoV-2-dependent epigenetic changes due to changes in their metabolic landscape. Additional experiments are necessary to elucidate this relevant aspect. In light of this consideration, a further question could be whether epigenetic changes might exist antecedently the first viral contact, persisting perhaps or worsening progressively up to the post-COVID19 period.

Several epigenetic phenomena have been associated with the SARS-CoV-2 infection (46), including the epigenetic regulation of ACE2 and IL-6, which has been reported to interest the development of a worse COVID19 due to excessive inflammation (47). Besides, the SARS-CoV-2 has been found to induce changes in DNA methylation, which affect the expression of immune response inhibitory genes that could, in part, contribute to the unfavorable progression of COVID-19 (48). Finally, it is noteworthy that a recently identified signature made of 44 variably methylated CpGs has been predictive of subjects at risk of developing worse symptoms after SARS-CoV-2 infection (49). Interestingly, none of these newly identified CpGs overlap with those involved in the DNAmAge prediction used in this (24) or other studies (29). Hypothetically, it might be possible that distinct signals are regulating the structure of the epigenomic regions determining a higher risk of developing a worse COVID19 syndrome and those associated with DNAmAge prediction.

Even though epigenetics might provide clinically relevant information about COVID19 (33) progression, no data is so far available about the involvement of epigenetic processes in the onset of the post-COVID19 syndrome or PPCS. Although none of the post-COVID survivors included in our study declared symptoms of PPCS, our evidence indicates the presence of changes in the methylation level of some CpGs associated with biological age calculation. This observation might reflect a more extensive phenomenon underlining unprecedented changes in the epigenome associated with the SARS-CoV-2 infection. A long-term follow-up of patients with an accelerated DeltaAge might be envisaged.

Telomere length is a marker of aging: progressive telomere shortening is a well-characterized phenomenon observed in older adults compared to the younger and attributed to the so-called telomeric attrition. This condition is worsened by the absence of telomerase activity physiologically silenced early post-natally and along adulthood (23). An accelerated TL shortening is a parameter associated with an increased risk of developing cardiovascular diseases and other disorders (50). In COVID19, patients bearing shorter telomeres in their peripheral leukocytes have been proposed to risk a worse prognosis (51). In the post-COVID19 group analyzed here, the average TL was 3,03 ± 2,39 Kb compared to the 10,67 ± 11,69 Kb of the control group (P<0.0001). As shown in Table 2, the chronological ages of the two cohorts were about comparable. Hence it is unlikely that the aging process could have been a determinant eliciting the difference. Accordingly, our results suggest that the observed TL shortening could be independent of DeltaAge (Figure 4B and C), indicating that the SARS-CoV-2 infection might directly contribute to telomere erosion in the blood cellular component.

ACE2 is a crucial component of the SARS-CoV-2 infection process. SARS-CoV-2 uses the ACE2 receptor to invade human alveolar epithelial cells and other cells, including cardiac fibroblasts (52). In infected individuals, ACE2 is often down-regulated due to the infection (11)(47). The enzyme is expressed in several tissues, including alveolar lung cells, gastrointestinal tissue, vascular cells, and the brain; however, it is relatively under-represented in circulating blood cells. In all cases studied, the total relative ACE2 mRNA level in the peripheral blood of non-COVID19 or post-COVID19 subjects was significantly lower than that of the MERS-CoV receptor DDP4. However, in the post-COVID19 group, the ACE2 mRNA expression was reduced significantly compared to controls, while DPP4 elicited similar expression levels in both groups. Interestingly, the accelerated DeltaAge, predominant in the younger Post-COVID19 survivors, significantly correlated with the lower ACE2 mRNA level, suggesting for an adverse effect of DNAmAge on ACE2 density in peripheral blood (Figure 5B and C).

The two groups considered in this study were not significantly different in terms of age, sex, and known clinical conditions before SARS-CoV-2 infection except for a relatively higher incidence of BMI>30 (15.3% vs. 9%) in the post-COVID19 population compared to controls as well as for a record of more frequent lung diseases (20.2% vs. 1.6%; see Table I). The origin of the persistent reduction in ACE2 expression in the post-COVID19 group remains unsolved, and a longitudinal study should be performed monitoring this parameter.

## Conclusions

This study has many significant limitations, including the small number of subjects investigated and the low number of CpGs considered. Although we used a valid forensic method to establish the biological age in the examined groups (24)(35) adopting other methods evaluating a large set of CpGs might be recommended (29)(53). However, the application of the latter procedures is limited by the elevated cost and relative complexity and may not be feasible at the laboratory level in many hospitals.

Nevertheless, it is shown here that individuals belonging to a group of COVID19 survivors had a significant acceleration of their biological age, occurring mainly in the younger individuals. This type of information has been correlated with TL shortening and the expression of ACE2 mRNA. It is too early to extrapolate whether relevant clinical indications may arise from this and other studies assessing the role of epigenetic changes in COVID19 syndrome (49) (48). However, a warning might be raised that sequelae of SARS-CoV-2 infection might rely on persistent epigenomic modifications, possibly underlying the presence of a COVID19 epigenetic memory. The epigenomic landscape of actual post-COVID19 survivors and in future COVID19 survivors from SARS-CoV-2 variants should then be taken under consideration to gain predictive prognostic insights and to monitor the response to treatment.

## Materials and methods

The samples have been classified into two groups: COVID19-free (n= 144), a heterogeneous group that includes healthies, cardiovascular diseases, and obstructive sleep apnea patients, while the post-COVID19 group includes all of the previous patients who had SARS-CoV-2 infection (n= 117). The clinical features of both populations are summarised in Table 2.

### DNA extraction from whole blood

Blood samples collected in EDTA and 200 ul have been used to perfom the extraction using QIAmp DNA Blood Mini Kit (Qiagen, cat. 55106) associated with automated system QIACube (Qiagen, cat. 9002160) according to manufacturer instructions. Then 2ul of DNA has been quantified with QIAxpert (Qiagen, cat. 9002340)

### Bisulfite Conversion

1ug of DNA has been used to convert with Epitect fast DNA bisulfite (Qiagen, cat. 59824) following the manufacturer instructions associated with RotorGene 2plex HRM (Qiagen, cat.9001560) and QIACube automated system. Then 2ul of converted DNA has been quantified with QIAxpert.

### Polymerase chain reactions for pyrosequencing

PCR reaction mixes were performed using PyroMark PCR kit (Qiagen, cat. 978103) following the manufacturer’s instructions. The sequences of primer used are available on Supplementary methods.

### Pyrosequencing

The amplicons have been sequenced in order to check the level of methylation in each CpG site. PyroMark Q24 Advanced Reagents (Qiagen, cat. 970902) have been loaded in the PyroMark Q24 Cartridge (Qiagen, cat.979202) following the manufacturer instructions, and 5ul of PCR product has been added to the reaction mix: Pyromark Binding Buffer (supplied in PyroMark Q24 Advanced Reagents kit), StreptavidinSepharose High performance (GE Healthcare, cat. GE17-5113-01) and H2O DNase/RNase free. Samples have been shacked at room temperature for 15 minutes at 1400rpm. Successively, the samples have been undergone to PyroMark Q24 Vacuum Station (Qiagen, cat. 9001515) procedure in which the target sequences have been purified and put into an annealing buffer containing the sequencing primer [0,375 uM]. The sequences of oligos are available in supplementary data. The plate containing the sequence to analyze and the primer has been heated at 80°C for 5minutes. Successively, the PyroMark Q24 Advanced (Qiagen, cat. 9001514) has been set to analyze the target sequences (Supplementary methods).

### DNAmAge estimation

Bekaert’s algorithm has been applied to estimate the biological age of the population (24) as reported in Daunay et al. (35): 26.44119-0.201902*ASPA-0.239205*EDARADD +0.0063745*ELOVL22+0.6352654*PDE4C

### Telomere length quantification

The chromosome end has been quantified by PCR Real-Time of Absolute Human Telomere Length Quantification qPCR Assay Kit (ScienCell, cat. 8918) following the manufacturer’s instructions.

### RNA extraction

According to manufacturer instruction, the total RNA has been isolated from whole blood using QIAmp RNA blood mini (Qiagen, cat. 52304) and automatized extractor QIACube. The RNA has been quantified with QIAxpert.

### cDNA synthesis and qPCR Real-Time

Omniscript RT kit (Qiagen, cat. 205113) has been used to convert total RNA into cDNA following the manufacturer’s instructions.

The qPCR RealTime has been performed on RotorGene 2plex HRM using the RT2 SYBR Green ROX FAST Mastermix (Qiagen, cat 330620). The sequences of primers are available on supplementary data. To perform the amplification; the machine has been set:

Initial denaturation: 95° C, 5 minutes;

Denaturation: 95° C, 15 seconds;

Annealing: 60° C, 30 seconds;

Elongation: 72° C, 30 seconds;

Final elongation: 72° C, 1 minute.

Denaturation, annealing, and elongation have been repeated for 45 times.

### Data analysis

All data has been analyzed with GraphPad Prism 8.4.3. Method to calculate the p-value: Two sided T-test.

## Data Availability

Ethical commettee ICS Maugeri document ID: 2543 CE

## List of abbreviations

(ARDS): adult respiratory distress syndrome
(ACE2): angiotensin-converting enzyme 2
(DNAmAge): biological age based on DNA methylation
(CVDs): cardiovascular diseases
(COVID19): coronavirus disease 19
(CpGs): Cytosine-Guanosine dinucleotides
(DPP-4): dipeptidyl-peptidase IV
(MERS): Middle East respiratory syndrome
(MERS-CoV): MERS coronavirus
(PPCS): persistent post-COVID19 syndrome
(RAS): renine-angiotensine system
(SARS): severe acute respiratory syndrome
(SARS-CoV-2): severe acute respiratory syndrome CoronaVirus 2
(TL): Telomere length
(T2DM): type 2 diabetes mellitus

## Declarations

This study have been approved by the Ethical committee of Istituti Clinici Scientifici Maugeri Spa SB di Pavia. Document: METH-COVID19 - Studio epigenetico sulle conseguenze dell’infezione da SarsCoV-2 (ID: 2543 CE)

## Author Contributions

Conceptualization,: AF, CG, TB and MTLR.; methodology: AM, VB, MGZ, SA; validation, CG AF, AP, FM, MM, MN.; formal analysis: AM, CG, MTLR, TB, SN; investigation: AM, VB, MGZ,SA, LF; resources: MN, VB, SN; data curation: AM; writing—original draft preparation: AM,CG; writing—review and editing: AF, TB, MTLR; supervision: CG; project administration: CG; funding acquisition: CG, MTLR.

## Acknowledgements

The authors would like to thanks Marta Lovagnini, Elena Robbi, Riccardo Sideri and Lorena Grano De Oro fpr valuable work and assistance providing patients information tracking and blood sample collection.

All authors have read and agreed to the published version of the manuscript.

